# Toward Non-Invasive Voice Restoration: A Deep Learning Approach Using Real-Time MRI

**DOI:** 10.1101/2025.08.22.25334256

**Authors:** Mohamad Saleh

## Abstract

Despite recent advances in brain–computer interfaces (BCIs) for speech restoration, existing systems remain invasive, costly, and inaccessible to individuals with congenital mutism or neurodegenerative disease. We present a proof-of-concept pipeline that synthesizes personalized speech directly from real-time magnetic resonance imaging (rtMRI) of the vocal tract, without requiring acoustic input. Segmented rtMRI frames are mapped to articulatory class representations using a Pix2Pix conditional GAN, which are then transformed into synthetic audio waveforms by a convolutional neural network modeling the articulatory-to-acoustic relationship. The outputs are rendered into audible form and evaluated with speaker-similarity metrics derived from Resemblyzer embeddings. While preliminary, our results suggest that even silent articulatory motion encodes sufficient information to approximate a speaker’s vocal characteristics, offering a non-invasive direction for future speech restoration in individuals who have lost or never developed voice.

## 1. Introduction

The ability to speak is a cornerstone of human communication and identity. Yet many individuals lose the capacity for intelligible speech due to advanced neurodegenerative diseases such as amyotrophic lateral sclerosis (ALS) [1], severe motor speech impairments following traumatic brain injury [2], or congenital conditions affecting vocal tract development [3]. Augmentative and alternative communication (AAC) devices can restore functional communication, but they often rely on synthetic voices that lack the emotional and personal nuances of the user [4]. Voice banking approaches allow individuals to preserve their vocal identity, but these require pre-recorded speech—an option unavailable to those who are congenitally mute or who experience sudden voice loss [5]. This gap highlights the urgent need for technologies that can generate a speaker’s unique vocal identity even in the absence of prior audio recordings.

Recent advances in brain–computer interfaces (BCIs) have enabled the decoding of neural activity into speech or text, offering a potential lifeline to patients with locked-in syndrome or advanced ALS [6,7]. While groundbreaking, these systems typically require invasive neurosurgical implantation, demand extensive training, and remain unsuitable for individuals who have never spoken, such as those born mute [8]. Non-invasive alternatives—such as video-based lip-reading models and silent speech interfaces—have shown promise in recognizing words from facial or articulatory movements [9,10]. However, these methods focus on reconstructing linguistic content rather than restoring a speaker’s unique vocal identity. Likewise, voice cloning systems depend on pre-recorded samples, making them inaccessible to individuals without prior vocal data [11].

These limitations highlight a critical gap in current speech restoration technologies: the need for methods capable of generating a natural-sounding voice purely from silent anatomical articulatory input.

To address this gap, we propose a proof-of-concept approach that leverages silent real-time magnetic resonance imaging (rtMRI) of the vocal tract to generate individualized voice characteristics. Specifically, we adapt the Pix2Pix generative adversarial network (GAN) [12] in a pipeline to translate segmented 2D rtMRI frames into synthetic audio representations. Unlike prior models aimed at speech recognition or linguistic decoding, our method does not seek to recover words or phonemes. Instead, it reconstructs an acoustic profile that approximates an individual’s vocal identity directly from articulatory anatomy. This introduces a new pathway toward identity-preserving voice synthesis that relies solely on silent articulatory motion, without requiring prior audio recordings or textual prompts.

In this proof-of-concept study, we train our system on paired data linking silent rtMRI segmentations of the vocal tract to articulatory class labels representing subject-specific configurations. A Pix2Pix GAN is used to map anatomical features to these identity-related representations, which are then transformed into synthetic audio by downstream neural models. We evaluate the generated voice profiles using Resemblyzer similarity metrics [13] and show that even a single silent articulatory posture can capture sufficient information to approximate aspects of a speaker’s unique vocal identity. These findings suggest a non-invasive pathway toward identity-preserving voice synthesis, with potential applications for individuals who have lost their voice or were born unable to speak.

## 2. Methods

### 2.1 Proposed Pipeline

We propose a multi-stage pipeline for synthesizing personalized speech audio from silent real-time MRI (rtMRI) frames of the vocal tract. The pipeline consists of three stages: rtMRI Preprocessing, Inference and Feature Mapping, and Audio Personalization as shown in Figure 1.

**Figure 1:**
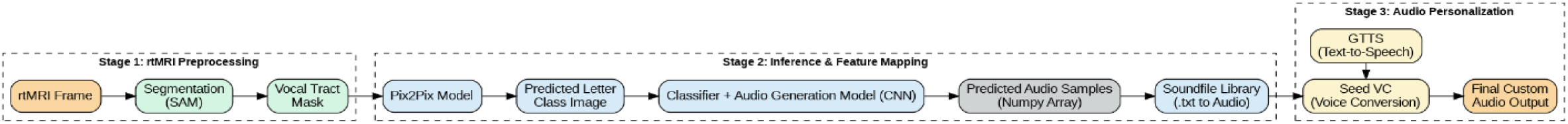
Overview of the proposed pipeline

**Stage 1: Preprocessing** – Input 2D rtMRI frames are segmented using the Segment Anything Model (SAM) [14] to isolate the vocal tract region, producing a mask that serves as input for subsequent stages.

**Stage 2: Inference and Feature Mapping** – The masked rtMRI frames are passed through a Pix2Pix image-to-image translation model [12] to generate predicted letter class images, which encode subject-specific vocal tract anatomy. These images are then classified using a combination of a visual classifier and a convolutional neural network (CNN) [15] to map each predicted class to its corresponding audio waveform. The resulting synthetic audio represents a low-level approximation of speech features and is rendered into audible form.

**Stage 3: Audio Personalization** – To achieve speaker-specific timbre, custom text input is converted to speech using the Google Text-to-Speech (gTTS) engine [16]. The output is then processed with the Seed Voice Conversion (Seed VC) model [17], which transforms the general TTS voice into a waveform reflecting the target speaker’s identity while preserving anatomical constraints derived from the rtMRI mask.

The modular architecture allows independent optimization of image translation, classification, and audio synthesis components. Combining CNN-based classification with Seed VC ensures that the final output is both anatomically grounded and perceptually natural.

### 2.2 Datasets

#### 2.2.1 Multispeaker RT-MRI Dataset

The multispeaker dataset of raw and reconstructed speech production real-time MRI video and 3D volumetric images [18] consists of 2D sagittal-view real-time MRI (rtMRI) videos synchronized with denoised audio recordings. The dataset includes recordings from 75 native American English speakers (male and female), each performing linguistically motivated speech tasks. The rtMRI videos have a spatial resolution of 84 × 84 pixels (2.4 × 2.4 mm²), acquired with a temporal resolution of 83.28 frames per second. The corresponding audio was low-pass filtered and resampled to a sampling rate of 20,000 Hz. For this study, data from 10 male speakers (sub006, sub0013, sub0015, sub0019, sub040, sub045, sub050, sub053, sub062, and sub072), aged between 23 and 36 years and all native speakers of American English, were selected based on age and linguistic background criteria. These subjects were considered for training purposes.

#### 2.2.2 USC-TIMIT rtMRI Dataset

The USC-TIMIT database [19] provides 2D mid-sagittal real-time MRI (rtMRI) videos synchronized with audio recordings of phonetically balanced sentences drawn from the MOCHA-TIMIT corpus. The dataset includes recordings from ten native speakers of American English (five male, five female), acquired at a spatial resolution of 68 × 68 pixels and a frame rate of 23.18 frames per second. The corresponding audio, recorded at 20,000 Hz, was contaminated with scanner noise, which was reduced using a custom adaptive filtering approach. In this study, data from three male speakers (M2 to M4) were used exclusively for testing, serving as unseen subjects in the evaluation phase.

### 2.3 Stages

#### 2.3.1 Stage 1: rtMRI Preprocessing

In the multispeaker rtMRI dataset, each of the ten participants was recorded articulating the sentence *“The North Wind and the Sun…”*. To enable anatomical comparison across speakers, we selected a single representative frame corresponding to the articulation of the word *“sun”*. This word was chosen due to its articulatory richness: the initial fricative /s/, the central vowel /ʌ/, and the final nasal /n/ each engage distinct and visible vocal tract structures, including the tongue, soft palate, and velum, which exhibit speaker-specific configurations [20].

To ensure consistency across speakers, we extracted the midpoint of the word *sun*, as it best captures the stable phase of articulation. The onset of a word often contains transient movement as the articulators shift from the previous sound, while the offset may reflect coarticulation with the following phoneme [21]. In contrast, the midpoint represents a relatively steady-state articulatory posture, where the tongue, lips, and velum are most fully engaged in producing the target sounds. Although some have argued that focusing on a single “magic moment” neglects the dynamic transitions that occur across a word [22], in this context the midpoint offers the clearest and most reproducible anchor point across subjects. This timing reduces variability due to frame selection and provides a consistent basis for visual and anatomical comparison, making it the most practical choice for our analysis.

Audio was extracted from each.mp4/.avi file using MoviePy [23] and transcribed with word-level timestamps using the Whisper speech recognition model [24]. The timestamp corresponding to the midpoint of “sun” was converted to a frame index based on the video’s frame rate, and the corresponding rtMRI frame was extracted using OpenCV [25] and saved for downstream analysis.

For the USC-TIMIT dataset, which includes recordings from multiple speakers reading phonetically balanced sentences, we selected subjects M2 through M4 and used sentences 001 to 005. Frame extraction was similarly performed by locating the “U” vowel in the word “sunshine”, which appears in sentence 004. The approximate timestamp for the vowel was estimated as one-sixth of the way through the word’s duration, and the corresponding frame was retrieved using the same Whisper–OpenCV pipeline.

To minimize anatomical variability and potential confounding effects on vocal tract shape, we selected only male, native American English speakers between the ages of 23 and 36 years from both datasets. Prior studies have shown that age, sex, and native language significantly influence vocal tract morphology and articulation patterns [26, 27, 28], motivating this constrained selection.

##### 2.3.1.1 Vocal Tract Segmentation with SAM-Med2D

To obtain a focused representation of the vocal tract from the extracted frames, we applied the SAM-Med2D model [29], a medical-domain adaptation of Meta AI’s Segment Anything Model (SAM), trained on the SA-Med2D-20M dataset [30]. Prior to inference, three manually placed point prompts were added to each frame: one at the lips, one at the posterior end of the vocal tract, and one at a midpoint along the tract as shown in Figure 2. These prompts guided the SAM-Med2D model to generate a smooth 2D mask of the vocal tract region.

**Figure 2:**
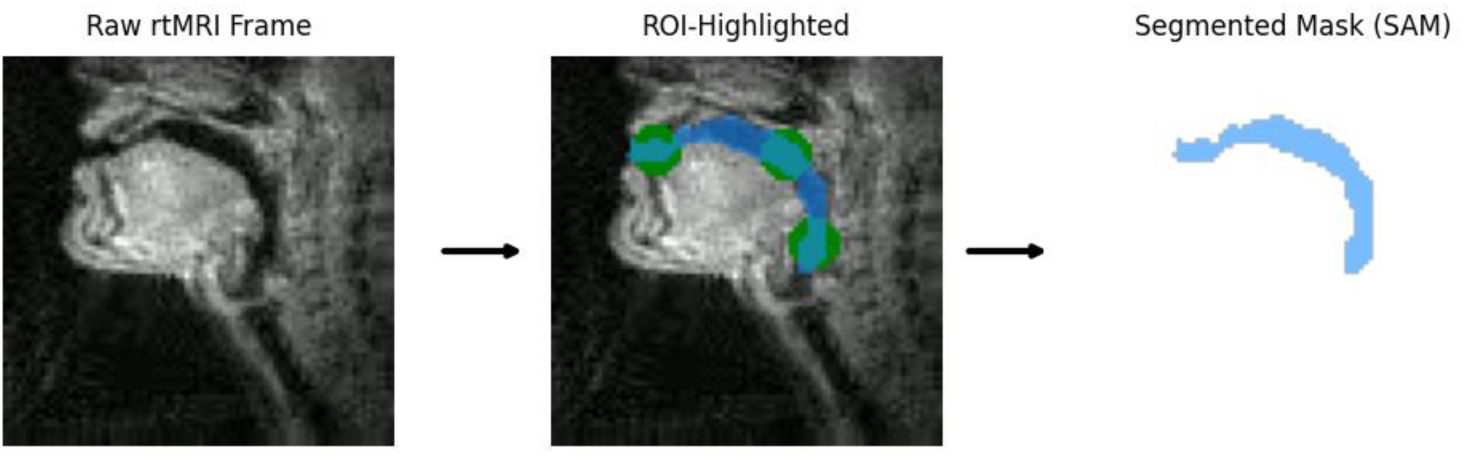
Segmentation of the Vocal tract with SAM-Med2D

This segmentation step was introduced for two primary reasons. First, rtMRI frames are inherently low-resolution and blurry [31], which could challenge models like Pix2Pix, making it difficult to learn shape-relevant patterns without noise. Second, isolating the vocal tract structure ensures that the model focuses on voice-relevant articulatory features, while minimizing distraction from irrelevant regions such as the head, jaw, or imaging artifacts.

##### 2.3.1.2 Subject Selection Criteria

To identify appropriate training subjects, we employed a two-step similarity-based selection process. First, we computed the Structural Similarity Index Measure (SSIM) [32] between generated vocal tract masks of candidate subjects and those of the held-out test subjects. SSIM is a perceptual metric that evaluates image similarity by comparing luminance, contrast, and structural information, yielding a score between 0 and 1, where higher values indicate greater structural resemblance. This allowed us to assess anatomical similarity of the vocal tract configurations.

In parallel, we used Resemblyzer [13], a neural network that encodes audio recordings into high-dimensional embeddings that capture speaker-specific characteristics. By computing cosine similarity between embeddings from candidate and test subjects, we obtained a measure of acoustic similarity, where scores closer to 1 indicate greater speaker identity resemblance. Subjects were considered for training only if they achieved a similarity score of 0.75 or higher for SSIM and 0.60 or higher for Resemblyzer relative to the test subjects.

Among the 10 subjects considered for training, we selected the three shown in Table 1. The computed similarity values are presented (SSIM and Resemblyzer scores), each comparing the three training subjects with the three test subjects (M2, M3, M4). Rankings guided the selection of training subjects to ensure strong articulatory and acoustic alignment. Subject 50 ranked first in SSIM and ninth in Resemblyzer similarity to M2, offering the best anatomical match and an acceptable acoustic similarity. Subject 6 ranked fifth in SSIM and first in Resemblyzer similarity to M3, achieving a strong vocal match with moderately similar anatomy. Subject 72 ranked first in both SSIM and Resemblyzer similarity to M4, indicating a near-perfect match across both modalities.

**Table 1:** SSIM and Resemblyzer scores, comparing the three training subjects (sub050,sub006,sub072) with the three test subjects (M2, M3, M4).

| **Subject** | **Compared to** | **SSIM** | **Resemblyzer Score** |
| --- | --- | --- | --- |
| M2 | Sub050 | 0.808 | 0.601 |
| M3 | Sub006 | 0.918 | 0.850 |
| M4 | Sub072 | 0.837 | 0.873 |

By selecting subjects that consistently ranked among the top in at least one metric while maintaining reasonable performance in the other, we ensured that the training data captured critical inter-subject similarities relevant to the model’s generalization. This approach reduced inter-subject variability and was critical for minimizing confounding effects in downstream tasks.

#### 2.3.2 Stage 2: Inference and Feature Mapping

##### 2.3.2.1 Brief Explanation of UNet, GANs, and Pix2Pix

UNet and Generative Adversarial Networks (GANs) are foundational concepts in image-to-image translation tasks. UNet [33] is a convolutional neural network architecture designed for semantic segmentation, which allows the model to produce high-quality pixel-wise predictions. Its encoder-decoder structure, with skip connections, enables the network to capture both low-and high-level features in an image. This makes it highly effective for tasks such as semantic segmentation, where precise pixel-level predictions are needed.

GANs [34] consist of two neural networks: a generator and a discriminator, which are trained in a competitive process. The generator aims to produce realistic data (such as images) to deceive the discriminator, while the discriminator’s goal is to differentiate between real and generated data. This adversarial training process enables GANs to generate realistic images in various domains, including image-to-image translation.

Pix2Pix [12] is a specific implementation of conditional GANs, where both the generator and discriminator are conditioned on input images. The generator learns to transform input images (such as low-resolution images or sketches) into realistic target images, while the discriminator ensures that the generated images are indistinguishable from real ones. This model has been widely used for tasks like image translation, super-resolution, and semantic segmentation, making it an ideal choice for generating corresponding anatomical labels from rtMRI frames.

##### 2.3.2.2 Speaker Grouping and Model Design Considerations

During the training phase, the three selected speakers (sub006, sub050, and sub072) were assigned to randomly-chosen identity-specific letter groups: “C”, “A”, and “L”, respectively. This grouping strategy reduced model complexity while preserving subject-specific vocal tract features. Each letter label served as a unique class representing the anatomical articulatory configuration of a specific speaker and was used as the target output image in the Pix2Pix training pipeline. Figure 3 illustrates the Pix2Pix model input-output relationship: a segmented rtMRI vocal tract mask is fed into the model, which generates an identity-specific letter image as output, representing the subject’s articulatory signature.

**Figure 3:**
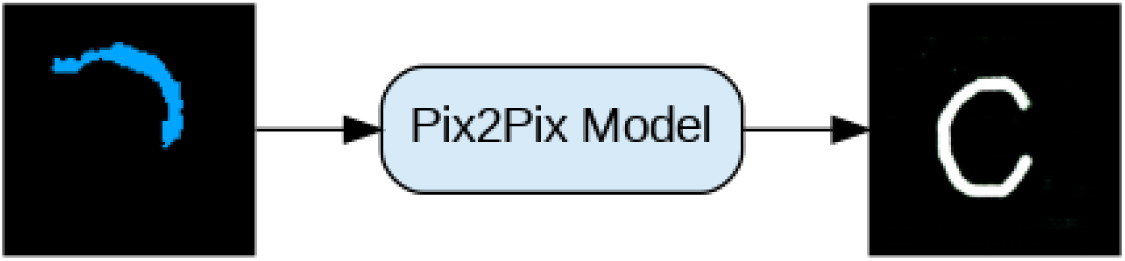
Pix2Pix model input-output relationship

To augment the training data and increase model robustness, we applied data augmentation [35, 36] by creating three color variants (green, light blue, and dark blue) of each subject’s vocal tract mask. This resulted in 100 copies of each subject, amounting to a total of 300 training samples. The generated color variations are shown in Figure 4, providing a visual representation of the augmentation process.

**Figure 4:**
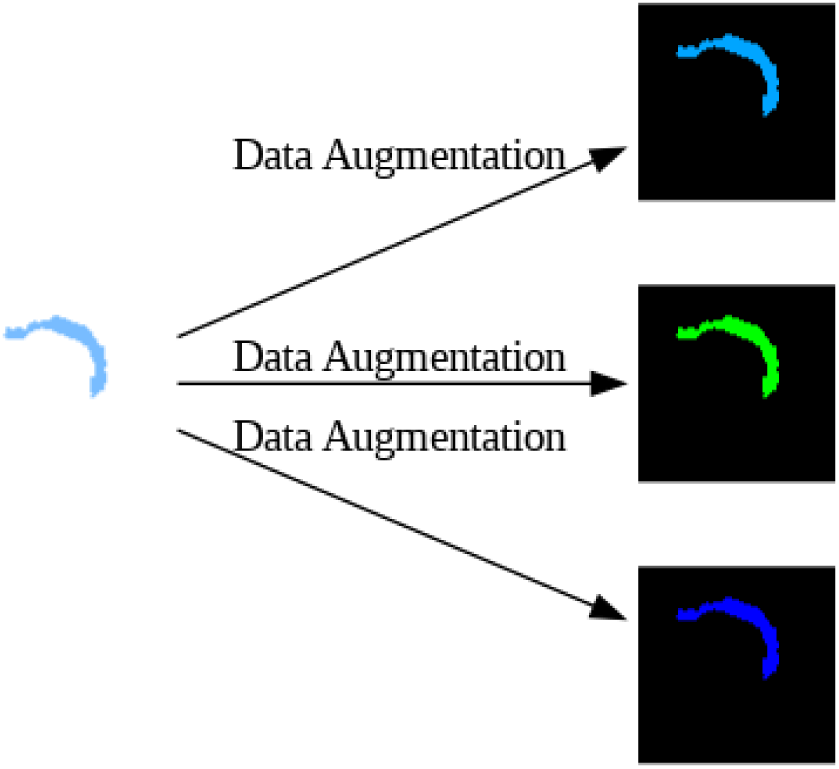
Data augmentation

Although Pix2Pix was initially considered for direct generation of spectrogram images from rtMRI frames, this approach was ultimately deemed suboptimal. The need to resize both inputs and outputs to 256×256 pixels led to a loss of fine-grained frequency and phase information, which is critical for high-fidelity waveform reconstruction [37]. Moreover, the generation of spectrograms requires significantly larger datasets and greater computational resources than were available in our setting. Consequently, Pix2Pix was limited to the task of generating subject-specific letter images from preprocessed and segmented vocal tract masks. A separate convolutional neural network (CNN) model was later trained to predict the corresponding NumPy array representations (used for speech reconstruction) from these generated letter images.

##### 2.3.2.3 Pix2Pix model training

Pix2Pix was trained with a learning rate of 0.00005, an L1 regularization weight of 100, and a batch size of 8. Training consisted of 200 epochs with a constant learning rate followed by 200 epochs with linear decay. Input images were resized and cropped during preprocessing, with no horizontal flipping applied. All training was performed on an NVIDIA T4 GPU, taking approximately 1.2 hours.

##### 2.3.2.4 Audio Generation and Voice Reconstruction

Rather than generating spectrograms directly, the Pix2Pix model was used exclusively to produce letter-class images corresponding to each speaker’s vocal tract anatomical identity. These generated letter images were first passed through a lightweight classifier that identified the represented letter (A, C, or L). The predicted letter class was then provided as input to a separate CNN model [15], which was designed to predict the corresponding audio samples in the form of numpy arrays [38]. The CNN model consisted of convolutional layers for feature extraction, followed by fully connected layers to map the features to the audio samples. For processing convenience and consistency with the vocoding pipeline, these arrays were exported as plain-text.txt files.

Figure 5 illustrates the overall pipeline, showing how the generated letter-class images from the Pix2Pix model are then processed by the classifier and CNN model to predict the corresponding audio.

**Figure 5:**
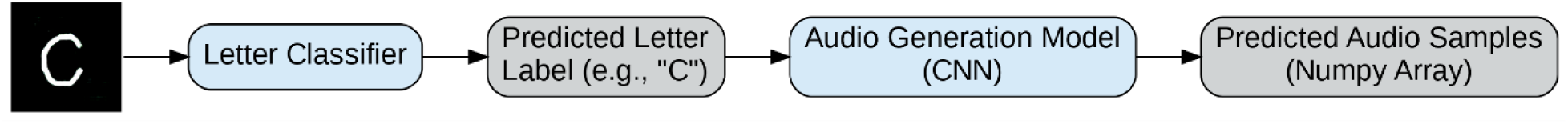
Letter Classification and Audio Samples Generation

###### 2.3.2.4.1 Letter Classification Model Configuration and Training

Following the generation of identity-specific letter images by the Pix2Pix model, a lightweight convolutional neural network (CNN) was employed to classify the predicted letters into one of three speaker-associated categories:’A’,’C’, or’L’. Each letter corresponds to a unique speaker whose vocal tract configuration was learned during training. The model was trained for 100 epochs on 30 letter PNGs generated by Pix2Pix, most of which were slightly distorted versions of the letters to allow for stronger classification. The classifier was trained on 128×128 grayscale letter images using a cross-entropy loss function and the Adam optimizer. The architecture consisted of two convolutional layers with ReLU activations, each followed by max pooling, and then a fully connected layer leading to a three-class softmax output. This module ensured accurate identification of the speaker label from the Pix2Pix output before passing it to the Audio Samples Generation model.

###### 2.3.2.4.2 Audio Samples Generation Model Configuration and Training

Once the letter identity was determined, the corresponding image was used as input to a regression model designed to predict audio samples in NumPy array format. This model was based on a convolutional neural network (CNN) for feature extraction, followed by fully connected layers to generate the audio representation. The input grayscale letter image was first flattened and passed through a linear layer to produce an embedded vector, which was then processed by the network and mapped to a 1D output corresponding to the audio samples. The model was trained for 30 epochs to generate 88,050 audio samples from the text files containing the NumPy arrays of the audios. It was trained using mean squared error (MSE) loss between the predicted and ground truth audio representations. The output arrays were saved as.txt files and later converted into audible speech using the SoundFile library [39]. This design allowed the system to separately control speaker identity (via classification) and speech content (via regression), while ensuring compatibility with downstream vocoding and evaluation tools.

#### 2.3.3 Stage 3: Audio Personalization

##### 2.3.3.1 Text-to-Speech Model Modification

For the audio generation component of this study, we selected Seed VC [17] primarily due to its high-quality voice cloning capabilities, making it well-suited for producing realistic audio. However, it is important to note that other Text-to-Speech (TTS) [40] systems with voice cloning capabilities could also have been used as alternatives to Seed VC. With the increasing accessibility of such models, systems like Tacotron 2 [41] and FastSpeech [42] can be substituted without significantly compromising performance. The key factor in model selection is the ability to synthesize speech that maintains high-quality audio characteristics.

To generate custom audio from text input, the model was integrated with Google Text-to-Speech (gTTS) [16]. The audio generated by gTTS served as the source audio for the model. gTTS was chosen for its clarity and neutral tone, which made it ideal for synthesis tasks.

The generated audio from the CNN-based regression model (trained to produce NumPy arrays corresponding to audio) served as the reference audio for comparison with the source audio. This allowed for generating custom speech that closely matched the target characteristics of the subject’s voice. The general pipeline for this process is illustrated in Figure 6 below.

**Figure 6:**
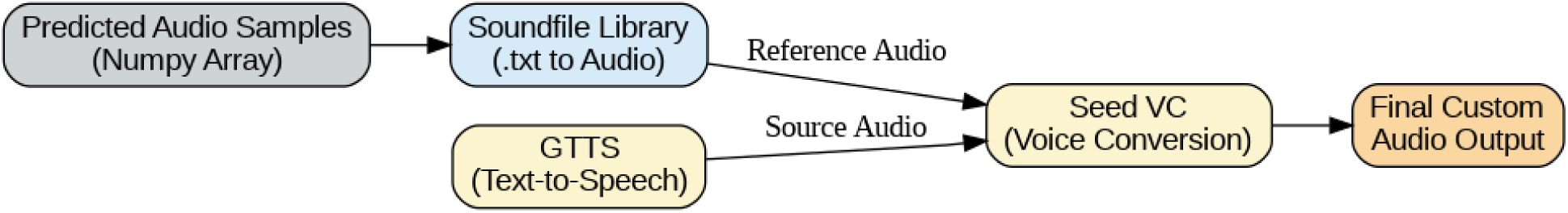
General pipeline for Audio synthesis and conversion.

### 2.4 Evaluation Metrics

Structural Similarity Index (SSIM) was used to evaluate how closely the anatomical-class letter images generated by the Pix2Pix model matched their ground truth counterparts. The similarity scores were then visualized using a bar graph to compare performance across the different vocal tract classes (’A’,’C’, and’L’).

Second, cosine similarity of speaker embeddings was employed to evaluate the quality of speaker identity preservation in the final synthesized audio. We used Resemblyzer [13], a deep speaker representation toolkit. Resemblyzer maps speech signals into a high-dimensional embedding space, where distances correspond to speaker similarity. The cosine similarity between the embeddings of the generated and reference audio is computed to evaluate how closely the generated voice resembles the target speaker.

Cosine similarity in Resemblyzer typically ranges from 0 (completely dissimilar) to 1 (perfect similarity). Values above 0.75 generally indicate strong speaker identity resemblance. This metric has been increasingly used in speaker identification and verification tasks and is known for its robustness across languages and noise conditions. In this study, we used Resemblyzer to extract embeddings from the Seed VC output and compare them to reference utterances of the target speakers in the unseen test set. We also applied it to evaluate the audio outputs generated by the letter classifier and Audio generation CNN model by comparing them to the original training subject audios, assessing how closely the predicted speech resembled the original voice in both training and testing contexts. This metric provided an objective measure of the effectiveness of the voice conversion module in personalizing synthesized speech.

## 3. Results and Discussion

Figure 7 shows SSIM scores for Pix2Pix-generated letter-class images compared to their ground truth equivalents. Scores ranged from 0.73 to 0.97, with the highest values for A1 and C1 (0.97), followed by L1 (0.93) and L7 (0.89). Lower scores (0.73–0.75) were concentrated in “C” class letters. Detailed inspection revealed that these reductions were primarily due to blurred or distorted Pix2Pix outputs for these classes. This effect is likely linked to the limited training data—only three subjects were used for model training—reducing exposure to the range of visual variations for certain letters and affecting output sharpness and structural accuracy.

**Figure 7:**
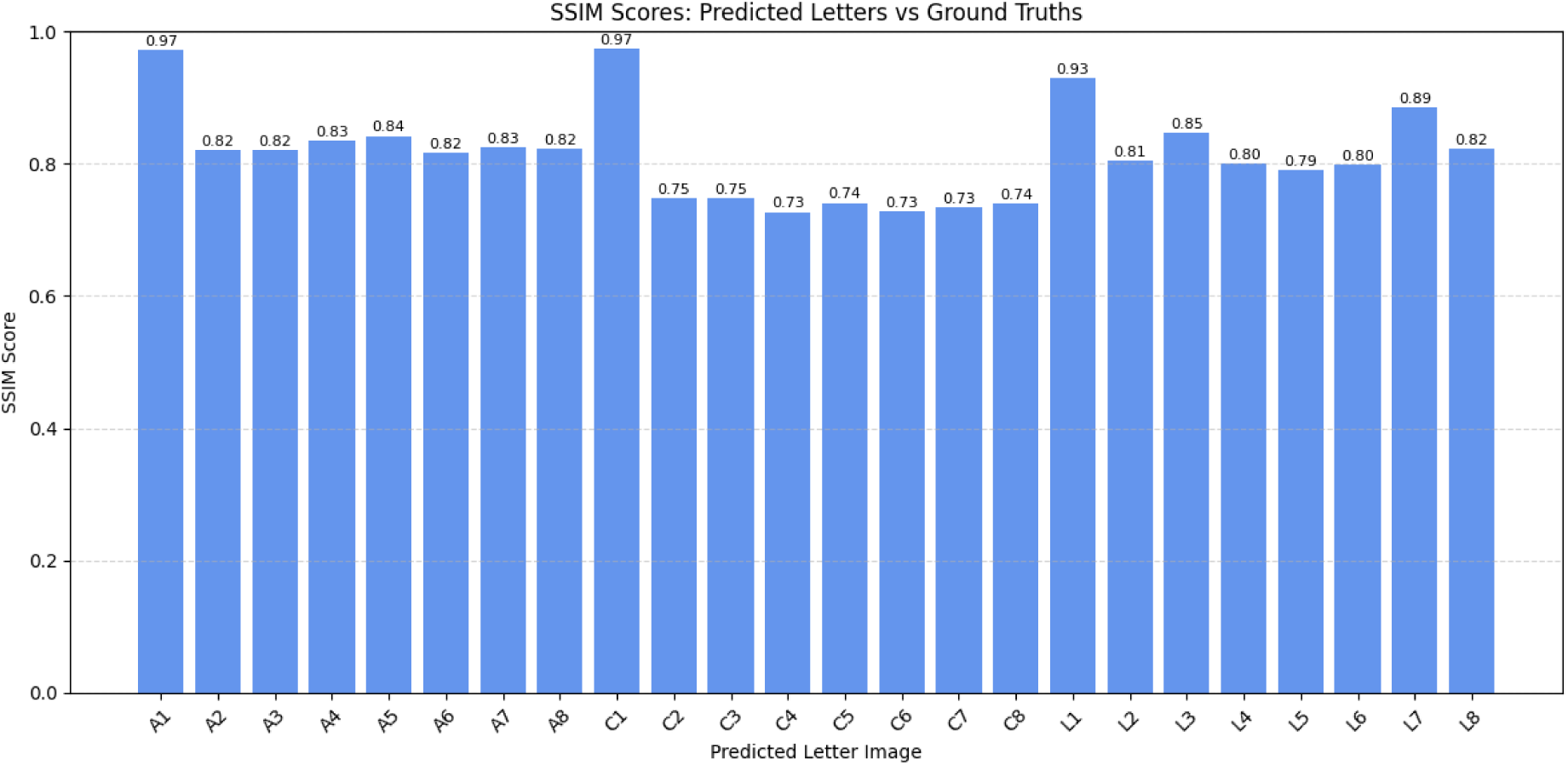
SSIM scores for Pix2Pix-generated letter-class images compared to their ground truth equivalents.

In addition to the quantitative evaluations, a qualitative test was performed to assess whether the Pix2Pix model relied on anatomical shape rather than color cues. Several segmented rtMRI masks were recolored with different hues while preserving their structural contours. Despite the altered colors, the model generated the correct letter-class images for these inputs, indicating that it had learned to differentiate classes based on anatomical features rather than relying on color consistency. Figure 8 shows an example of a recolored mask for an unseen subject M2 and the corresponding Pix2Pix output, demonstrating accurate class generation under altered visual conditions.

**Figure 8.**
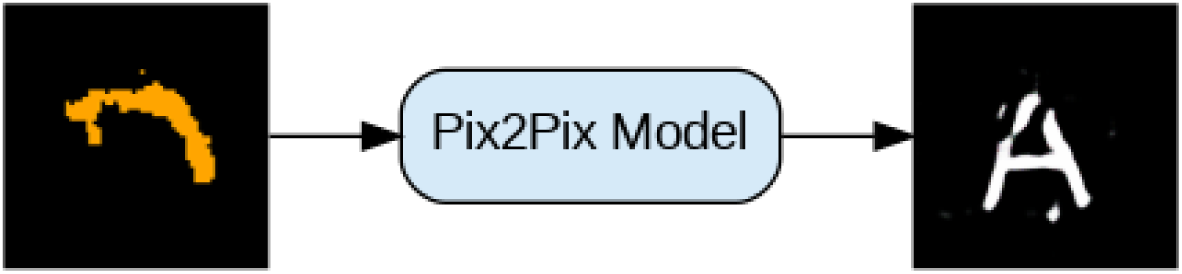
Example of a recolored segmented rtMRI mask for an unseen subject M2 (left) and its corresponding Pix2Pix-generated letter-class image (right). The correct class was produced despite the altered input color, indicating that the model differentiates vocal tract masks based on structural features rather than color information.

Table 2 presents Resemblyzer similarity scores comparing predicted audio generated by the proposed pipeline to the original audio of both the same and unseen subjects. In this context, a segmented single-frame rtMRI mask is first processed by Pix2Pix to produce a letter-class image representing the anatomical class. This image is then classified into its correct letter label, which is mapped to pre-generated audio samples in the subject’s voice. The predicted audio in Table 2 is therefore the result of mapping classified letters from visual data back to voice. Scores ranged from 0.540 (M2 vs. Sub050) to 0.940 (Sub006), with an overall average of 0.775. Higher similarity values were generally observed for same-subject comparisons, indicating that the mapping preserved much of the speaker-specific identity. Lower scores, especially in cross-subject comparisons, suggest a natural decrease in acoustic resemblance when predictions were evaluated against speakers not included in training.

**Table 2:** Resemblyzer similarity scores comparing predicted audio generated by the proposed pipeline to the original audio of both the same and unseen subjects.

| Original Audio | Predicted Audio | Resemblyzer Score |
| --- | --- | --- |
| Sub050 | Sub050 | 0.890 |
| Sub006 | Sub006 | 0.940 |
| Sub072 | Sub072 | 0.840 |
| M2 | Sub050 | 0.540 |
| M3 | Sub006 | 0.750 |
| M4 | Sub072 | 0.690 |

Table 3 compares custom audio—produced by the Seed VC model using gTTS-synthesized neutral speech as source audio and the predicted audio as the reference—to the original audio of both same and unseen subjects. In this case, the gTTS output serves as a neutral sample, while the predicted audio (from the letter-to-voice mapping) is used to condition the Seed VC model to match the target speaker. Scores ranged from 0.610 (M2 vs. Sub050) to 0.860 (Sub006), with an average of 0.763. The close performance between same and unseen subjects suggests that Seed VC is robust in adapting neutral synthetic speech into the target speaker’s style, even when the conditioning audio is generated rather than recorded.

**Table 3:**
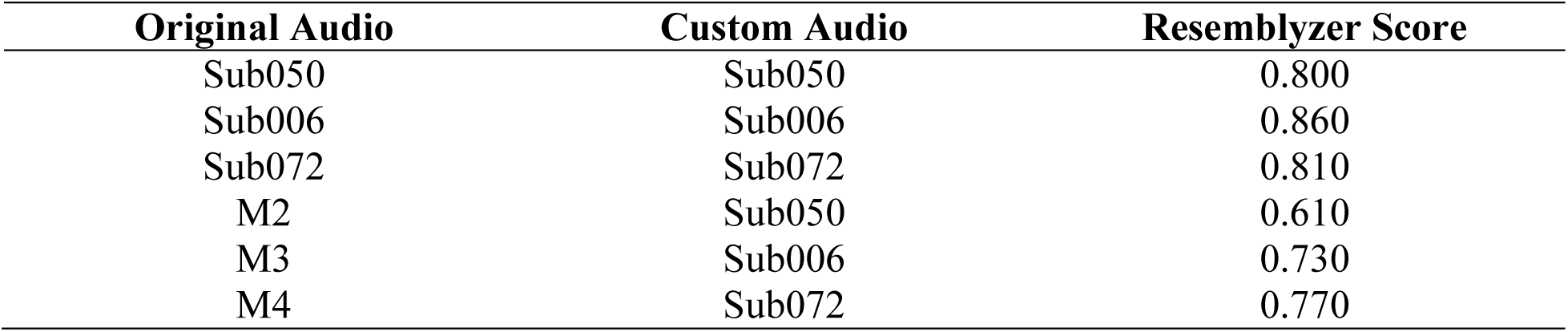
Resemblyzer similarity scores comparing custom audio generated by the Seed VC model to the original audio of both the same and unseen subjects.

Overall, the results show that the proposed pipeline:

1. Generated letter-class images from segmented rtMRI masks with high structural accuracy for many classes.
2. Correctly classified these images and mapped them to subject-specific audio with strong same-subject and unseen-subject similarity to original recordings.
3. Enabled effective personalization of neutral gTTS speech into target-speaker style via Seed VC, even with generated source audio.

These findings demonstrate the feasibility of using anatomical-class classification and letter-to-voice mapping for silent speech synthesis while highlighting the need for increased training data diversity, especially for classes with lower SSIM scores.

## 4. Conclusion

This work introduces an exploratory, proof-of-concept pipeline for synthesizing speech directly from rtMRI of the vocal tract using deep generative models. By linking anatomical representations with acoustic outputs, the approach demonstrates the potential for approximating personalized speech without reliance on recorded audio. While preliminary, these findings highlight a non-invasive direction for future speech restoration technologies. Taken together, this study provides an early step toward non-invasive, data-driven speech synthesis for individuals who are unable to speak. Future research should focus on larger and more diverse datasets, automated segmentation, and prospective testing in clinical populations to better establish feasibility.

## 5. Limitations

This study should be interpreted within its constraints. The dataset was limited to 10 participants, and only a subset of subjects showing strong correlations between mask similarity and voice similarity were included, limiting generalizability. Segmentation of rtMRI frames using the SAM model was performed manually, so slight deviations in SSIM scores are expected. In addition, the seed voice conversion (VC) model produces minimal run-to-run variation in voice similarity metrics, meaning small fluctuations may reflect model behavior rather than true performance differences. Finally, the approach has not yet been validated in individuals who are mute or have speech disorders; any extension to these populations remains hypothetical.

## Data Availability

All data produced in the present study are available upon reasonable request to the authors.

https://figshare.com/articles/dataset/A_multispeaker_dataset_of_raw_and_reconstructed_speech_production_real-time_MRI_video_and_3D_volumetric_images/13725546

https://sail.usc.edu/span/usc-timit/index.html

## References

[1] Duffy, J. R. (2012). Motor speech disorders: Substrates, differential diagnosis, and management. Elsevier Health Sciences.

[2] McHenry, M. (2000). Acoustic characteristics of voice after severe traumatic brain injury. The Laryngoscope, 110(7), 1157–1161.

[3] Gray, S. D., Smith, M. E., & Schneider, H. (1996). Voice disorders in children. Pediatric Clinics of North America, 43(6), 1357–1384.

[4] Pullin, G., & Hennig, S. (2015). 17 ways to say yes: Toward nuanced tone of voice in AAC and speech technology. Augmentative and Alternative Communication, 31(2), 170–180.

[5] Cave, R., & Bloch, S. (2021). Voice banking for people living with motor neurone disease: Views and expectations. International Journal of Language & Communication Disorders, 56(1), 116–129.

[6] Littlejohn, K. T., Cho, C. J., Liu, J. R., Silva, A. B., Yu, B., Anderson, V. R.,… & Anumanchipalli, G. K. (2025). A streaming brain-to-voice neuroprosthesis to restore naturalistic communication. Nature neuroscience, 1-11.

[7] Willett, F. R., Kunz, E. M., Fan, C., Avansino, D. T., Wilson, G. H., Choi, E. Y.,… & Henderson, J. M. (2023). A high-performance speech neuroprosthesis. Nature, 620(7976), 1031–1036.

[8] Séguin, P., Maby, E., Fouillen, M., Otman, A., Luauté, J., Giraux, P.,… & Mattout, J. (2024). The challenge of controlling an auditory BCI in the case of severe motor disability. Journal of NeuroEngineering and Rehabilitation, 21(1), 9.

[9] Pham, D. N., & Rahne, T. (2025). Development and evaluation of a deep learning algorithm for German word recognition from lip movements. arXiv preprint arXiv:2504.15792.

[10] Liu, Q., Ge, M., & Li, H. (2024). Intelligent event-based lip reading word classification with spiking neural networks using spatio-temporal attention features and triplet loss. Information Sciences, 675, 120660.

[11] Neekhara, P., Li, J., & Ginsburg, B. (2021). Adapting TTS models for new speakers using transfer learning. arXiv preprint arXiv:2110.05798.

[12] Isola, P., Zhu, J. Y., Zhou, T., & Efros, A. A. (2017). Image-to-image translation with conditional adversarial networks. In Proceedings of the IEEE conference on computer vision and pattern recognition (pp. 1125–1134).

[13] Resemble AI. (2020, November). Resemblyzer: A Python package to analyze and compare voices with deep learning [Computer software]. GitHub. https://github.com/resemble-ai/Resemblyzer

[14] Ravi, N., Gabeur, V., Hu, Y. T., Hu, R., Ryali, C., Ma, T.,… & Feichtenhofer, C. (2024). Sam 2: Segment anything in images and videos. arXiv preprint arXiv:2408.00714.

[15] O’shea, K., & Nash, R. (2015). An introduction to convolutional neural networks. arXiv preprint arXiv:1511.08458.

[16] Gobinathan, A., Hashim, N. M. Z., & Omar, A. (2024). TTS-UTeM: Text to Speech Transformation Application. APS, 44.

[17] Liu, S. (2024). Zero-shot voice conversion with diffusion transformers. arXiv preprint arXiv:2411.09943.

[18] Lim, Y., Toutios, A., Bliesener, Y., Tian, Y., Lingala, S. G., Vaz, C.,… & Narayanan, S.S. (2021). A multispeaker dataset of raw and reconstructed speech production real-time MRI video and 3D volumetric images. Scientific data, 8(1), 187.

[19] Narayanan, S., Toutios, A., Ramanarayanan, V., Lammert, A., Kim, J., Lee, S.,… & Proctor, M. (2014). Real-time magnetic resonance imaging and electromagnetic articulography database for speech production research (TC). The Journal of the Acoustical Society of America, 136(3), 1307–1311.

[20] Proctor, M., Bresch, E., Byrd, D., Nayak, K., & Narayanan, S. (2013). Paralinguistic mechanisms of production in human “beatboxing”: A real-time magnetic resonance imaging study. The Journal of the Acoustical Society of America, 133(2), 1043–1054.

[21] Fletcher, A. R., McAuliffe, M. J., Lansford, K. L., & Liss, J. M. (2017). Assessing vowel centralization in dysarthria: A comparison of methods. *Journal of Speech*, Language, and Hearing Research, 60(2), 341–354.

[22] Carignan, C., Hoole, P., Kunay, E., Pouplier, M., Joseph, A., Voit, D.,… & Harrington, J. (2020). Analyzing speech in both time and space: Generalized additive mixed models can uncover systematic patterns of variation in vocal tract shape in real-time MRI. Laboratory Phonology: Journal of the Association for Laboratory Phonology, 11.

[23] Zulko. (2017). MoviePy: A software library for video editing [Computer software]. GitHub. https://github.com/Zulko/moviepy.

[24] Radford, A., Kim, J. W., Xu, T., Brockman, G., McLeavey, C., & Sutskever, I. (2023, July). Robust speech recognition via large-scale weak supervision. In International conference on machine learning (pp. 28492–28518). PMLR.

[25] Bradski, G. (2000). The opencv library. Dr. Dobb’s Journal: Software Tools for the Professional Programmer, 25(11), 120–123.

[26] Vorperian, H. K., Kent, R. D., Gentry, L. R., & Yandell, B. S. (1999). Magnetic resonance imaging procedures to study the concurrent anatomic development of vocal tract structures: Preliminary results. International Journal of Pediatric Otorhinolaryngology, 49(3), 197–206.

[27] Markova, D., Richer, L., Pangelinan, M., Schwartz, D. H., Leonard, G., Perron, M.,… & Paus, T. (2016). Age-and sex-related variations in vocal-tract morphology and voice acoustics during adolescence. Hormones and behavior, 81, 84–96.

[28] Heyne, M., Derrick, D., & Al-Tamimi, J. (2019). Native language influence on brass instrument performance: An application of generalized additive mixed models (GAMMs) to midsagittal ultrasound images of the tongue. Frontiers in psychology, 10, 2597.

[29] Cheng, D., Qin, Z., Jiang, Z., Zhang, S., Lao, Q., & Li, K. (2023). Sam on medical images: A comprehensive study on three prompt modes. arXiv preprint arXiv:2305.00035.

[30] Ye, J., Cheng, J., Chen, J., Deng, Z., Li, T., Wang, H.,… & Qiao, Y. (2023). Sa-med2d-20m dataset: Segment anything in 2d medical imaging with 20 million masks. arXiv preprint arXiv:2311.11969.

[31] Otani, Y., Sawada, S., Ohmura, H., & Katsurada, K. (2023, August). Speech synthesis from articulatory movements recorded by real-time mri. In Proc. Interspeech (Vol. 2023, pp. 127–131).

[32] Wang, Z., Bovik, A. C., Sheikh, H. R., & Simoncelli, E. P. (2004). Image quality assessment: from error visibility to structural similarity. IEEE transactions on image processing, 13(4), 600–612.

[33] Ronneberger, O., Fischer, P., & Brox, T. (2015, October). U-net: Convolutional networks for biomedical image segmentation. In International Conference on Medical image computing and computer-assisted intervention (pp. 234–241). Cham: Springer international publishing.

[34] Goodfellow, I. J., Pouget-Abadie, J., Mirza, M., Xu, B., Warde-Farley, D., Ozair, S.,… & Bengio, Y. (2014). Generative adversarial nets. Advances in neural information processing systems, 27.

[35] Perez, L., & Wang, J. (2017). The effectiveness of data augmentation in image classification using deep learning. arXiv preprint arXiv:1712.04621.

[36] Kim, E. K., Lee, H., Kim, J. Y., & Kim, S. (2020). Data augmentation method by applying color perturbation of inverse PSNR and geometric transformations for object recognition based on deep learning. Applied Sciences, 10(11), 3755.

[37] Tamiti, T. I., Joshi, B., Hasan, R., Hasan, R., Athay, T., Mamun, N., & Barua, A. (2025). A High-Fidelity Speech Super Resolution Network using a Complex Global Attention Module with Spectro-Temporal Loss. arXiv preprint arXiv:2507.00229.

[38] Harris, C. R., Millman, K. J., Van Der Walt, S. J., Gommers, R., Virtanen, P., Cournapeau, D.,… & Oliphant, T. E. (2020). Array programming with NumPy. nature, 585(7825), 357–362.

[39] Bechtold, B. (2015). SoundFile: An audio library based on libsndfile, CFFI, and NumPy [Computer software]. GitHub. https://github.com/bastibe/python-soundfile

[40] Hasanabadi, M. R. (2023). An overview of text-to-speech systems and media applications. arXiv preprint arXiv:2310.14301.

[41] Shen, J., Pang, R., Weiss, R. J., Schuster, M., Jaitly, N., Yang, Z.,… & Wu, Y. (2018, April). Natural tts synthesis by conditioning wavenet on mel spectrogram predictions. In 2018 IEEE international conference on acoustics, speech and signal processing (ICASSP) (pp. 4779–4783). IEEE.

[42] Ren, Y., Ruan, Y., Tan, X., Qin, T., Zhao, S., Zhao, Z., & Liu, T. Y. (2019). Fastspeech: Fast, robust and controllable text to speech. Advances in neural information processing systems, 32.

